# Photon-Counting Detector Computed Tomography Angiography to assess intracranial stents and flowdiverters - in vivo study comprising ultra-high resolution spectral reconstructions

**DOI:** 10.1101/2024.08.06.24311513

**Authors:** Frederic De Beukelaer, Sophie De Beukelaer, Laura L Wuyts, Omid Nikoubashman, Mohammed El Halal, Iliana Kantzeli, Martin Wiesmann, Hani Ridwan, Charlotte S. Weyland

**Affiliations:** Department of Neuroradiology, University hospital RWTH Aachen, Aachen, Germany; Department of Neurology, Inselspital, University hospital Bern, Bern, Switzerland; Department of Radiology, AZ Sint-Lucas, Ghent Belgium

**Keywords:** Computed Tomography Angiography, Stent, Cerebral Arterial Diseases, Photon-Counting

## Abstract

**BACKGROUND AND PURPOSE:** Neuroimaging of intracranial vessels with implanted stents (ICS) and flowdiverters (FD) is limited by artifacts. Photon-Counting-Detector-Computed Tomography (PCD-CT) is characterized by a higher resolution. The purpose of this study was to assess the image quality of ultra-high-resolution (UHR) PCD-CT-Angiography (PCD-CTA) and spectral reconstructions to define the best imaging parameters for the evaluation of vessel visibility in ICS and FD.

**MATERIALS AND METHODS:** Retrospective analysis of consecutive patients with implanted ICS or FD, who received a PCD-CTA between April 2023 and March 2024. Polyenergetic (PE), virtual monoenergetic imaging (VMI), pure lumen (PL) and iodine (I) reconstructions with different kiloelectron volt (keV) levels (keV 40, 60 and 80) and reconstruction kernels (Body vascular kernel (Bv) 48, Bv56, Bv64, Bv72, Bv76) were acquired to evaluate image quality and assessed by 2 independent radiologists using a 5-point Likert scale and regions of interest (ROI). The different kernels, keV and the optimized spectral reconstructions were compared in descriptive analysis.

**RESULTS:** In total, 12 patients with 9 FDs and 6 ICSs were analyzed. In terms of quantitative image quality, sharper kernels as Bv64 and Bv72 yielded increased image noise, and decreased signal to noise (SNR) and contrast to noise ratio (CNR) compared to the smoothest kernel Bv48, (p<0.01). Among the different keV levels and kernels, readers selected the 40 keV level (p<0.01) and sharper kernels (in the majority of cases Bv72) as the best to visualize the in-stent vessel lumen. Assessing the different spectral reconstructions virtual monoenergetic and iodine reconstructions proved to be best to evaluate in-stent vessel lumen (p<0.01).

**CONCLUSIONS:** Our preliminary study suggests that PCD-CTA and spectral reconstructions with sharper reconstruction kernels and a low keV level of 40 seem to be beneficial to achieve optimal image quality for the evaluation of ICS and FD. Iodine and virtual monoenergetic reconstructions were superior to pure lumen and polyenergetic reconstructions to evaluate in-stent vessel lumen.

**Key points:** PCD-CTA imaging quality can be advanced by image post processing with spectral reconstructions and certain reconstruction kernels.

Intracranial in-stent vessel lumen visibility significantly improves using sharp reconstruction kernels (Bv72) and low keV level in high-resolution mode in photon-counting detector computed tomography. Regarding the different optimized spectral reconstructions virtual monoenergetic and iodine reconstructions proved to be best to evaluate in-stent vessel lumen.

## Introduction

Photon-Counting Detector CT (PCD-CT) offers high spatial resolution and various spectral reconstruction possibilities to improve neurovascular imaging where Energy-integrating-Detector (EID)-CT is limited [1-3].

Phantom studies for PCD-CT reported promising results for stent assessment with reduced artifacts[4]. In comparison to EID-CT, two studies describing a total of seventeen patients, found higher iodine contrast attenuation (20.8%) with virtual monoenergetic imaging (VMI) reconstructions and less beam-hardening in regions with extensive bone involvement (C2 segment of the carotid artery) [5,6].

In vivo studies comparing different reconstruction kernels identified Bv48 in the cerebrovascular angiography module as optimal for assessing intracranial aneurysms [7].

Cardiovascular angiographies have identified higher kernels as better to evaluate coronary arteries: The vessel lumen of coronary arteries after stent implantation was best evaluated at a sharper reconstruction kernel (Body vascular kernel (Bv) 60 and 72) [8].

Studies reporting on imaging post processing in PCD-CT involving spectral reconstructions are scarce in neurovascular imaging. In vitro studies suggest that for monoenergetic reconstructions the optimal energy threshold is between 40 and 60 kiloelectron volt (keV) to identify iodine [9].

Intracranial stents (ICS) and flowdiverters (FD) have become widely used in the past years to treat intracranial stenosis and wide-necked aneurysms. PCD-CT-Angiography (PCD-CTA) using different keV levels and reconstruction kernels might offer the possibility to assess patients with implanted intracranial stents or flowdiverters using non-invasive imaging as compared to diagnostic subtraction angiography (DSA) for patient follow-up.

We aimed to assess the imaging post-processing possibilities of PCD-CTA to find the best reconstruction kernels and keV-levels to assess lumen visibility and artifacts of different ICS and FD.

## Methods

This is a single-center retrospective analysis of consecutively treated patients, who were treated for intracranial atherosclerotic disease with symptomatic, high-grade stenosis or wide-necked aneurysms using ICS or flowdiverters FD between April 2023 and May 2024.

### Assessment of CT data

Patients underwent a CTA centered on the implanted device acquired on a clinical first-generation PCD-CT scanner (NAEOTOM Alpha, Siemens Healthineers, Erlangen, Germany) operated in ultra-high-resolution (UHR) mode. Polyenergetic (PE), virtual monoenergetic imaging (VMI), pure lumen (PL) and iodine (I) reconstructions with different kiloelectron volt (keV) levels (keV 40, 60 and 80 were acquired. PCD-CT image acquisition and image post processing details involving spectral reconstructions specifics can be found in the electronic supplemental data.

Two radiologists with 9 years (blinded for peer review) and 10 years (blinded for peer review) of experience assessed all images independently (workstation: syngo.via, version VB10A, Siemens Healthineers). Regions of interest (ROIs) were manually placed on each PCD-CTA reconstruction. A total of five manually drawn ROI for each reconstruction were placed: One in the proximal parent vessel, one in the proximal end of the ICS or FD and one in the middle of the ICS or FD. To assess the CNR one ROI was placed in the air, rostral to the forehead and one in the superficial temporal muscle. ROIs were copied and then pasted into the other three reconstruction imaging, allowing to place the ROIs at the exact same location with the exact same size on every PCD-CTA reconstruction.

The size of the ROIs was as large as possible, ensuring that only the lumen of the artery was measured, carefully avoiding the stent struts. Signal was defined as the average density (Hounsfield units, HU) and noise as the standard deviation (SD) of ROI density. For the calculation of the signal-to-noise ratio (SNR) and contrast-to-noise ratio (CNR), the muscle density in the superficial temporal muscle (signal_muscle_) and the SD of the air (SD_air_) adjacent to the rostral neurocranium were measured. SNR and CNR were calculated as follows:

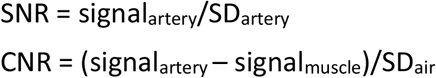

The two readers assessed image quality of the in-stent lumen visibility at the proximal end of the stent and in the stent using a 5-point Likert scale, ranging from 1 (“non-diagnostic”) to 5 (“excellent”).

### Ethics

This study was approved by the local ethics committee (BLINDED FOR PEER REVIEW). The study fulfills the STROBE criteria, and the study was conducted following the International Declaration of Helsinki[10].

### Statistics

IBM SPSS Statistics software (version 28.0) was used for statistical analysis.

Normal distribution for descriptive analysis was tested using the Shapiro Wilk test. Quantitative variables were expressed as mean ± standard deviation (SD) or median and interquartile ranges (IQR). Scores from the qualitative image quality analysis were pooled across readers.

Friedman’s test was used to compare the different kernels in each reconstruction (I, PL, PE, VMI) with pairwise comparisons. In case of the VMI and PL reconstructions, the three keV levels were compared for each kernel before using the test. Friedman’s test was used to compare the Likert values of the different optimized spectral reconstructions (I, PL, PE, VMI).

Interobserver agreement was expressed in Cohen’s kappa (κ) value and interpreted as follows: ≤ 0.20 as none, 0.21–0.40 as fair, 0.41–0.60 as moderate, 0.61–0.81 as substantial, and ≥ 0.82 as almost perfect agreement. All p-values were corrected for multiple testing using the Bonferroni-correction. Reliability of the Likert scale was tested using Cronbach’s alpha test.

A 95% confidence interval range (CI) was calculated and expressed for the results of all diagnostic accuracy tests. A two-tailed p-value of < 0.05 was considered statistically significant.

All in all, we evaluated 480 reconstructions and placed 2400 ROIs. Results were visualized with ggplot2 and Likert packages within R Software. (The R Project for Statistical Computing (r-project.org)) [11].

### Results

The study population included 12 patients, of which 11 were women. Mean age at the time of the PCD-CTA scan was 55.5 (± 12.7) years. The indications of the PCD-CTA included aneurysm follow-up or postinterventional control. 8 (67%) patients had intracranial aneurysms at following locations: 5 (62%) intracranial segments of the internal carotid artery, 1 (13%) V4-segment of the right vertebral artery, 1 (13%) posterior communicating artery. 7 (87%) aneurysms were treated with a total of 9 FD (2 p48 HPC and 7 p64 MW HPC, Phenox®, Germany). 1 (13%) aneurysm of the distal M1-segment of the right middle cerebral artery was treated with stent assisted coiling (Acclinoflex®, Acandis GmbH, Pforzheim, Germany). Mean vessel diameter was 2,7 mm ± 0.65.

4 stenosis of intracranial vessels were treated with a total of 4 stents (2 CREDO® and 1 Acclino® (Acandis GmbH, Pforzheim, Germany), 1 Neuroform Atlas® (Stryker Neurovascular, Fremont, California, USA) at following locations: 1 (25%) basilar artery, 1 (25%) medial cerebral artery, 1 (25% anterior cerebral artery), 1 (25%) internal carotid artery.

Patient population characteristics can be found in Table 1.

**Table 1.** Patient characteristics.

|  |  |  |
| --- | --- | --- |
| Age (y)* |  | 55.5 (± 12.7) |
| Sex (female/male) |  | 11 / 1 |
| Aneurysm locations | Internal cerebral artery | 5 |
|  | Medial cerebral artery | 1 (13) |
|  | Anterior cerebral artery | 0 |
|  | Posterior communicating artery | 1 (13) |
|  | Vertebral artery | 1 (13) |
|  | Basilar artery | 0 |
| Stenosis locations | Internal cerebral artery | 1 (25) |
|  | Medial cerebral artery | 1 (25) |
|  | Anterior cerebral artery | 1 (25) |
|  | Posterior communicating artery | 0 |
|  | Vertebral artery | 0 |
|  | Basilar artery | 1 (25) |
| Flowdiverter type | p48 HPC | 2 |
|  | p64 HPC | 6 |
| Stent type | Acclino | 3 |
|  | Credo | 2 |
|  | Neuroform atlas | 1 |
| In-Stent vessel diameter (mm)* |  | 2.7 (± 0.67) |

Note. — Except where indicated, data are numbers of participants, with percentages in parentheses.
Abbreviations: y=years,;\* Data are means ± standard deviations

### Quantitative Analysis – PCD-CTA Reconstructions

The results of the quantitative and qualitative analysis are displayed in Figure 1-3.

**Figure 1:**
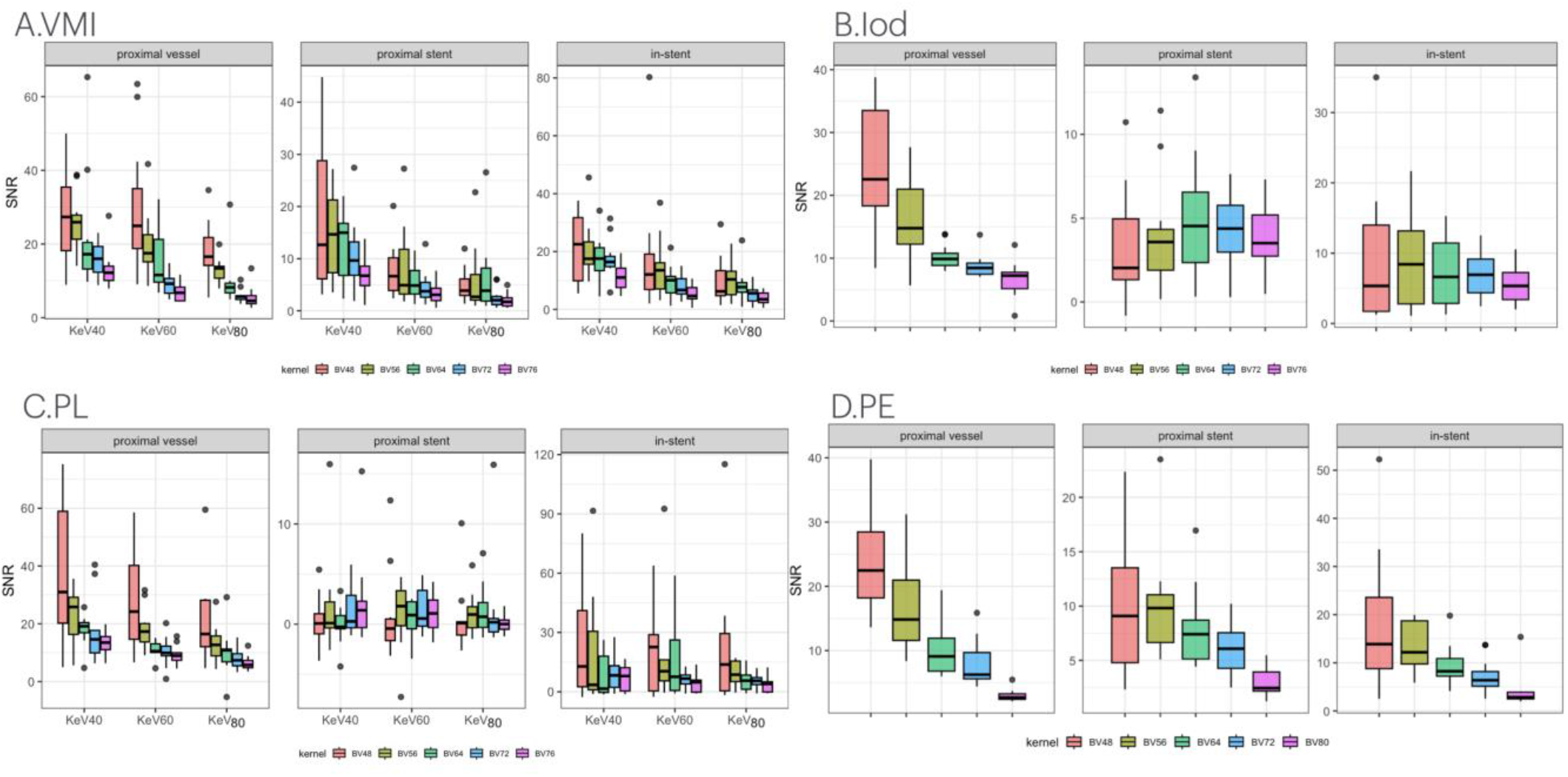
Boxplot Distribution of Signal-to-Noise Ratio (SNR) in Photo-Counting-Detector CT-Angiography (PCD-CTA) images for 5 reconstruction kernels (Bv48, Bv56, Bv64, Bv72, Bv76 (Bv80 for PE)) in different spectral reconstructions (A-D). For virtual mono energetic (VMI; see A) and pure lumen (PL; see C), distributions are assessed at 3 keV levels (40, 60 and 80keV). Abbreviations: VMI= virtual monoenergetic imaging, PL=pure lumen, PE=polyenergetic, Bv= Body Vascular. KeV= kiloelectrovolt. The lower and upper quartile, representing observations outside the 9 –91 percentile range. The diagram also shows the median and mean observation for a particular keV or kernel. Data falling outside the Q1 – Q3 range are plotted as outliers of the data.

**Figure 2.**
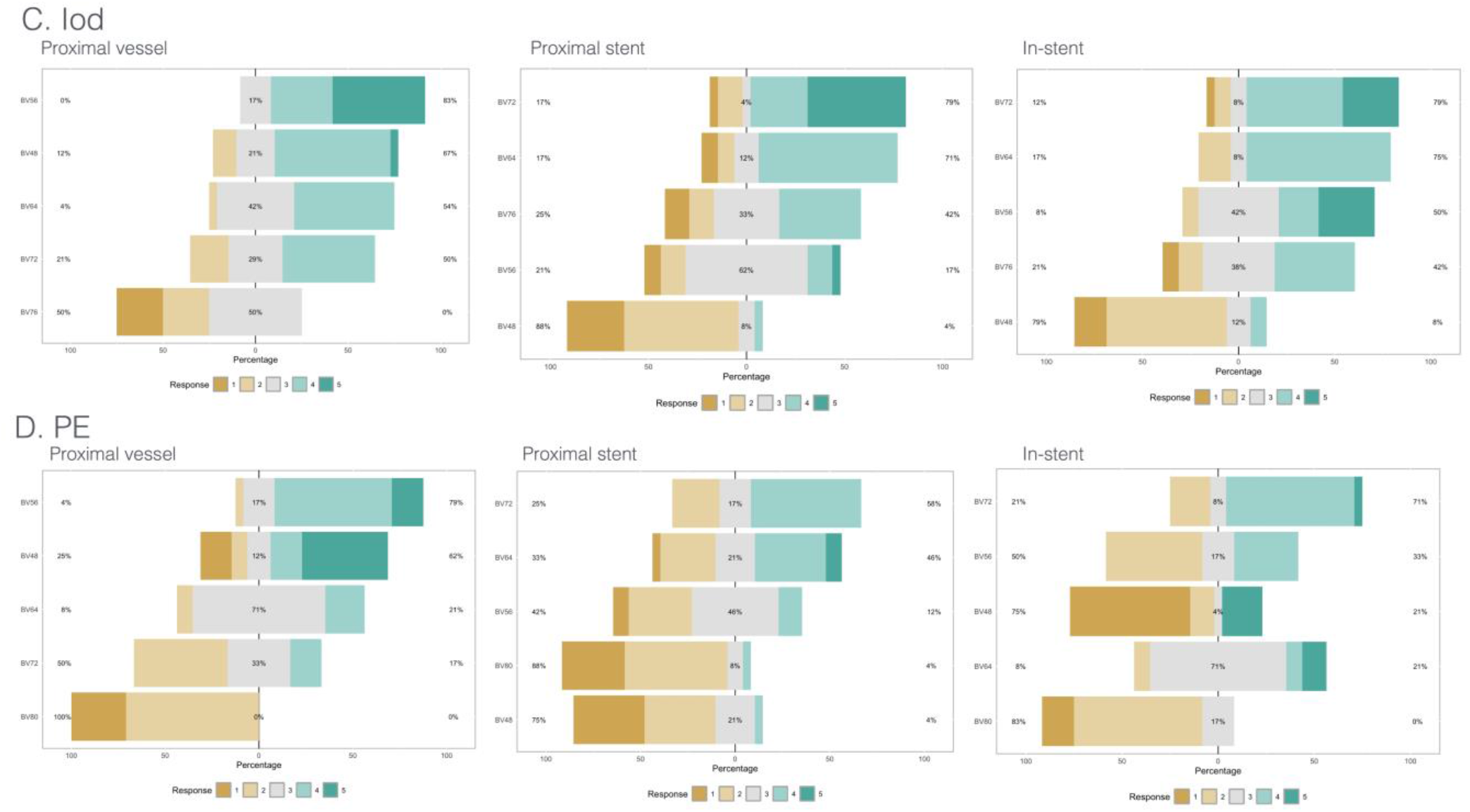
Qualitative image quality scores from the PCD-CTA images (Bv48, Bv56, Bv64, Bv72, Bv76 (for Iod reconstructions (C)) and Bv80 (for polyenergetic reconstructions (PE)). Stacked bar charts show pooled percentages of two raters, at three different localsations, i.e. proximal vessel, proximal stent, in-stent. Interpretation of scores: 5 = excellent image quality, 4 = good image quality, 3 = acceptable image quality, 2 = barely satisfactory image quality, 1 = unacceptable image quality. (Bv = Body vascular kerneel, Iod= Iodine reconstructions, PE = polyenergetic reconstructions, PCD-CTA = Photon-counting detector CT-angiography)

**Figure 3.**
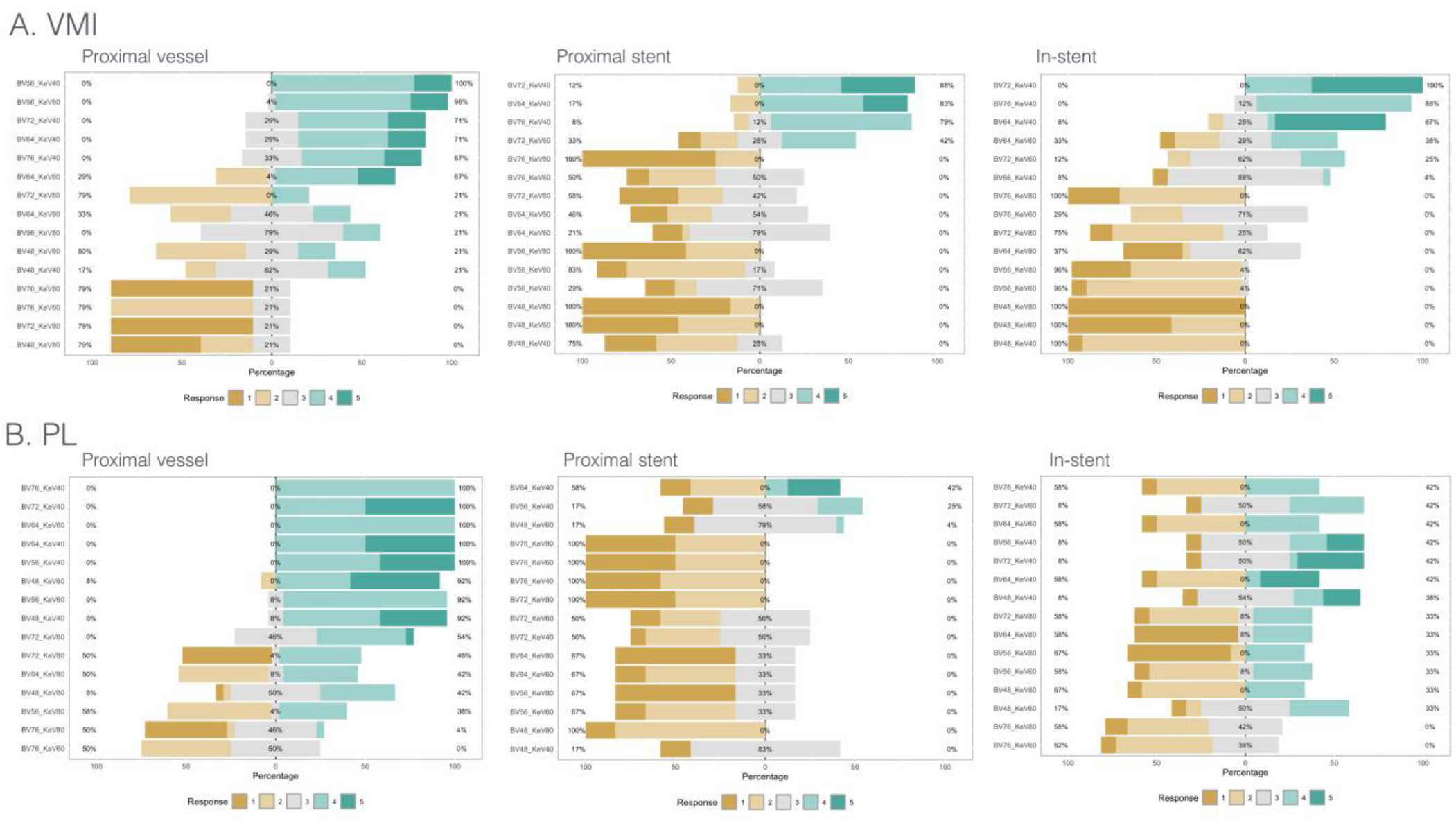
Qualitative image quality scores from the PCD-CTA images according to different spectral reconstructions, here VMI (A) and PL (B) of different kernels (Bv48, Bv56, Bv64, Bv72, Bv76) and keV levels (40, 60, 80). Stacked bar charts show pooled percentages of two raters, at three different localsations, i.e. proximal vessel, proximal stent, in-stent. Interpretation of scores: 5 = excellent image quality, 4 = good image quality, 3 = acceptable image quality, 2 = barely satisfactory image quality, 1 = unacceptable image quality. (Bv = Body vascular, keV= kiloelectron Volt, PL = Pure Lumen, PCD-CTA = Photon-counting detector CT-Angiography, VMI= Virtual monoenergetic imaging).

Detailed information concerning the individual SNR/CNR values for the various reconstructions can be found in the electronic supplemental data.

Virtual monoenergetic imaging reconstructions (VMI)

For (VMI) an incremental decrease of SNR and CNR was found across the range of kernel and keV levels:

1. In VMI, the lowest keV-level (40) provided the highest SNRs and CNRs, mostly with significant differences compared to the highest keV level (80). As an example, SNR values for the Bv48 kernel was higher at 40 keV than at 80 keV in the intracranial parent vessel (mean ± SD: keV 40: 27.5 ± 11.7 vs. keV 80: 18.1 ± 7.5, p = 0.01), at the proximal ICS/FD end (mean ± SD; keV 40: 46.9 ± 12.8 vs. keV 80: 4.8 ± 3.1, p < 0.01) and in-stent vessel lumen (mean ± SD; keV 40: 21.7 ± 11.3 vs.keV 80: 9.8 ± 8.1, p < 0.01).

2. In VMI, transitioning from the smoothest (Bv48) to the sharpest (Bv76) kernel, the smoothest kernel provided the highest SNRs and CNRs. As an example, SNR values for Bv48 and Bv76 kernels for 40keV were in the intracranial parent vessel (mean ± SD: 27.5 ± 11.7 vs. 12.2 ± 4.7, p<0.01), at the proximal ICS/FD end (mean ± SD: 46.9 ± 12.8 and 7.1 ± 3.5, p<0.01) and in-stent vessel lumen (mean ± SD: 21.7 ± 11 and 10.9 ± 4.7, p<0.01).

### Pure Lumen reconstructions (PL)

For pure lumen (PL) reconstructions, an incremental decrease of SNRs and CNRs was found across the range of kernels and keV levels. This reconstruction was most affected by artifacts of the implanted devices, especially stent markers.

For the intracranial parent vessel, the lowest keV-level (40) provided the highest SNR and CNR, mostly with significant differences compared to the highest keV-level (80). As an example, SNR values for the Bv48 kernel for 40 keV and 80 keV were in the intracranial parent vessel (mean ± SD: 39.5 ± 25.7 and 23.8 ± 21.7, p = 0.02).

Transitioning from the smoothest (Bv48) to the sharpest (Bv76) kernel, the sharpest kernel provided the highest SNR and CNR. As an example, SNR values for the Bv48 and Bv76 kernels for 40keV were in the intracranial parent vessel (mean ± SD: 39.5 ± 25.7 and 14.9 ± 3.2, p<0.01).

Significant differences of SNR and CNR values for the in-stent vessel lumen could not be detected across different kernels and keV levels.

### Iodine reconstructions

For iodine reconstruction an incremental decrease in SNR was found for the intracranial vessels across the range of kernels, transitioning from the smoothest (Bv48) to the sharpest (Bv76) kernel in the intracranial parent vessel (from 24.6 ± 9.9 to 6.6 ± 2.8, p<.01). In case of in-stent vessel lumen no significant difference in SNR could be found.

### Polyenergetic reconstructions

For polyenergetic reconstructions an incremental decrease of SNR and CNR was found across the range of kernels and keV-levels: Transitioning from the smoothest (Bv48) to the sharpest (Bv80) kernel, the smoothest kernel provided the highest SNRs and CNRs. As an example, SNR values for the Bv48 and Bv80 kernels were in the intracranial parent vessel (mean ± SD: 24.6 ± 8.6 and 2.7 ± 1.1, p<0.01), at the proximal ICS/FD end (mean ± SD: 10.4 ± 6.4 and 3.1 ± 1.3, p<0.01) and in-stent vessel lumen (mean ± SD: 18.9 ± 14.3 and 4.2±3.7, p<.01).

### Qualitative Analysis

Substantial agreement (α = .72) was found between the two readers for qualitative ratings using the 5-point Likert-type scale. The internal consistency of the test, measured by Cronbach’s alpha, was excellent (.89).

For VMI, images reconstructed with 40 keV achieved the highest score (p<.01). For the parent vessel images reconstructed with Bv56 kernel achieved the highest scores (p<.01).

For the in-stent vessel lumen, Bv64, Bv72, Bv76 achieved higher scores than Bv56 and Bv48. Out of the three kernels, the two readers of the study favored the Bv72 kernel for the visualization of in-stent vessel lumen.

For pure lumen reconstructions, images reconstructed with the 40keV achieved the highest scores, however, for the parent vessel as well as for the in-stent vessel lumen, there was no preferred reconstruction.

For iodine reconstructions, images reconstructed with the Bv56 kernel achieved higher assessment scores for the parent vessel. For evaluation of the in-stent vessel lumen, Bv72 achieved higher scores than Bv56 and Bv48. Out of the three kernels, the two readers of the study favored the Bv72 kernel.

For the polyenergetic reconstructions, the parent vessel images reconstructed with the kernels Bv48 and Bv56 kernels achieved the highest score. However, imaging with Bv48/Bv56 and Bv64 kernels was comparable. Out of the three kernels, the two readers of the study favored the Bv56 kernel for the visualization of the parent vessel.

For the in-stent vessel lumen, the Bv64 and Bv72 kernels achieved higher scores than Bv80, Bv56 and Bv48 kernels. However, only the pairwise comparisons of Bv64 and Bv72 kernels with the Bv80 kernel was significant (p<.01)/(p<.01).

When reviewing the PCD-CTA images in consensus reading after blinded assessment, the Bv72 kernel was selected as the most suitable for the evaluation of in-stent vessel lumen.

When assessing the best spectral reconstructions to evaluate in-stent vessel lumen, pairwise comparisons identified virtual monoenergetic images at 40 keV and iodine reconstructions as best to evaluate in-stent vessel lumen (p<0.01).

## DISCUSSION

The present study evaluates the objective and subjective image quality of PCD-CTA for intracranial vessels after intracranial stent or flowdiverter implantation using different levels of kernel sharpness and keV. Our results highlight the significance of sharper kernels in enhancing the in-stent vessel lumen. The use of the Bv72 kernel across spectral reconstructions and, additionally, a low keV level (40 keV) for virtual monoenergetic reconstructions significantly increased vessel visibility.

Studies on kernel optimization for in-stent vessel lumen of intracranial arteries have been limited to phantom studies. Based on their objective and subjective image quality results, the authors recommended the use of Bv60 for spectral PCD-CTA [4].

This study represents a systematic approach for neurovascular imaging applications across different kernel reconstructions and spectral reconstructions using PCD-CTA.

Our quantitative analysis showed that image noise increases by utilizing sharper kernels, resulting in a continuous decrease in SNR and CNR, even for a quantum iterative reconstruction (QIR)-level of three.

The qualitative image analysis demonstrated a clear preference of the readers for sharper kernels. A similar observation was published recently: Qualitative assessment differed from the quantitative assessment [7]. We share the authors’ opinion, that for a comprehensive evaluation of image quality, both qualitative and quantitative assessments should be considered, as they provide complementary information.

While SNR and CNR allow a reproducible, quantitative evaluation for assessing image quality, qualitative evaluation allows to integrate several features simultaneously, that may be difficult to quantify.

The Bv64 and Bv72 kernels both delivered superior qualitative image quality. However, the observers favored the Bv72 kernel (at a keV-level of 40) for evaluating the in-stent vessel lumen.

While digital subtraction angiography remains the “gold standard” method for the evaluation of intracranial aneurysms, PCD-CTA has the potential to offer less invasive imaging for FD and ICS evaluation.

Our preliminary results may provide a valuable reference to exploit the full potential of the ultra-high-resolution mode and spectral reconstructions of PCD-CTA for neurovascular imaging.

The presented results have the following limitations:

First, this preliminary study included a relatively small number (12) of patients from a single center, which limits the generalizability of our results, and implanted FD and ICS were of limited variability.

Second, only one type of kernel (Body vascular kernel) was evaluated on PCD-CTA. Third, ROIs were placed manually by a single observer, which could introduce the risk of measurement bias.

## CONCLUSION

Our preliminary study suggests that PCD-CTA and spectral reconstructions with sharper reconstruction kernels and a low keV level of 40 seem to be beneficial to achieve optimal image quality for the evaluation of intracranial vessels after intracranial stent or flowdiverter implantation. Iodine and virtual monoenergetic reconstructions were superior to pure lumen and polyenergetic reconstructions to evaluate in-stent vessel lumen.

## Data Availability

All data produced in the present study are available upon reasonable request to the authors

